# COVID-19 Vaccine Acceptance Among Healthcare Workers in a United States Medical Center

**DOI:** 10.1101/2021.04.29.21256186

**Authors:** Michelle H. Moniz, Courtney Townsel, Abram L. Wagner, Brian J. Zikmund-Fisher, Sarah Hawley, Charley Jiang, Molly J. Stout

**Affiliations:** University of Michigan Department of Obstetrics and Gynecology; University of Michigan Institute for Healthcare Policy and Innovation; University of Michigan Department of Epidemiology; University of Michigan Department of Health Behavior and Health Education; University of Michigan Department of Internal Medicine; University of Michigan Center for Bioethics and Social Sciences in Medicine

## Abstract

**Background:** The Centers for Disease Control and Prevention prioritized healthcare personnel for the first phase of COVID-19 vaccination in the United States to keep critical healthcare infrastructure open and functioning, but vaccine hesitancy may limit vaccine uptake.

**Objective:** To evaluate vaccine intentions among healthcare workers eligible for COVID-19 vaccination and explore differences by sociodemographic and occupational characteristics.

**Design:** From February 1-15, 2021, we conducted a cross-sectional, opt-in online survey at a Midwest U.S. academic healthcare center that began vaccinating employees in December 2020.

**Participants:** The entire employee workforce of the study site was eligible.

**Main Measures:** COVID-19 vaccination intention, categorized as Received/Scheduled/ASAP, Not Now, and Not Ever. Logistic regression models to assess the relationship between demographic and occupational characteristics and intention to receive COVID-19 vaccination.

**Key Results:** Most participants (n=11,387, of 39,259 individual and group email accounts invited) had received or were scheduled to receive the COVID-19 vaccine (n=9081, 79.8%) or planned to receive it as soon as possible (n=546, 4.8%), while fewer were hesitant (Not Now, n=954, 8.4%; Not Ever, n=369, 3.2%). In multivariable logistic regression models predicting vaccine intention, physicians (aOR 22.2, 9.1-54.3), trainees (aOR 5.9, 3.0-11.4), and nurse practitioners/nurse midwives/physician assistants (aOR 1.9, 1.2-3.0) were significantly more likely to demonstrate vaccine acceptance, compared to nurses, whereas other clinical staff were significantly less likely (aOR 0.8, 0.6-0.9). Prior infection with COVID-19, gender, race/ethnicity, and age were all significantly associated with vaccine intention. Overall, 29.6% reported at least one concern about COVID-19 vaccination.

**Conclusions:** In a large, diverse sample of healthcare workers, over 11% delayed COVID-19 vaccination when it was available to them, with notable variation in vaccine hesitancy across professional roles and demographic groups. Our findings suggest immediate opportunities to empathetically engage those with COVID-19 vaccine concerns and optimize vaccine coverage across our healthcare system.

## INTRODUCTION

COVID-19 vaccines hold promise to minimize hospitalizations and deaths due to SARS-CoV-2, which has claimed the lives of more than 512,000 Americans to date.^1^ The Federal Drug Administration (FDA) issued Emergency Use Authorizations (EUAs) for the Pfizer-BioNTech and Moderna COVID-19 vaccines in December 2020 and for the Johnson & Johnson vaccine in February 2020.^2,3^ Recognizing that vaccine supply would be limited initially, the Centers for Disease Control and Prevention^4^ prioritized healthcare personnel for the first phase of vaccination in the United States to keep critical healthcare infrastructure open and functioning. By the end of December 2020, healthcare systems across the country began offering COVID-19 vaccines to employees.

However, access does not guarantee uptake,^5^ and it remains unclear to what degree healthcare workers will accept the COVID-19 vaccine as it is made available to them. In one study of long-term care facility staff, only 38% of eligible workers received COVID-19 vaccination in December 2020-January 2021.^6^ A survey of 1000 healthcare workers in January 2021 documented widespread vaccine hesitancy, with 1 in 4 respondents anticipating declining COVID-19 vaccination.^7,8^ Moreover, prior work during the H1N1 influenza pandemic demonstrated variation in vaccine acceptance across roles of healthcare employees.^9^ It is currently unclear how COVID-19 vaccine acceptance and hesitancy might vary across the diverse healthcare employee ecosystem, which includes clinicians, other staff not providing clinical care but with clinical contact (e.g., unit clerks, hospitality staff, security staff), administrators, and researchers.

It is critical to address this knowledge gap to inform ongoing and future vaccine distribution efforts. When a healthcare worker declines a COVID-19 vaccine, it affects the herd immunity of the healthcare workforce and potentially the safety of patients and communities that workforce serves. Healthcare workers also constitute a trusted voice that influences others—vaccine hesitancy in this group could undermine efforts to widely vaccinate populations and achieve herd immunity, as has been modeled for seasonal influenza.^10^ Better understanding of healthcare workers’ attitudes and beliefs about COVID-19 vaccination could identify opportunities to create more effective vaccine messaging and policies and ultimately promote more widespread vaccine coverage and improved population health outcomes.

Our objective was to evaluate vaccine intentions and attitudes among a diverse sample of medical center employees eligible for COVID-19 vaccination and to explore variations in vaccine beliefs among employees with different occupational and demographic characteristics.

## METHODS

### Study population

We conducted a cross-sectional, opt-in online survey of the entire employee workforce at an academic healthcare center in the U.S. Midwest in from February 1-15, 2021. COVID-19 vaccine administration started at the healthcare center on December 14, 2020. All employees at the center were eligible for vaccination. None were required to be vaccinated.

### Survey Administration

We administered an online survey in English to all medical center employees via Qualtrics. All employees with a medical center email address were eligible to participate, including frontline clinicians (e.g., nurses, physicians, respiratory therapists, medical assistants); other staff not providing clinical care but with clinical contact (e.g., unit clerks, environmental services staff); trainees, administrators, and other faculty and staff with no clinical contact. On February 1, 2021, the President of the health system sent an email invitation to participate in the survey using a global user email list that includes 39,259 individuals, as well as groups, in the healthcare system. A reminder email was sent nine days later. The survey was open for a total of 14 days.

### Survey Instrument

We developed the survey instrument through a rapid, iterative, and collaborative process involving all members of our interprofessional study team, which included clinical experts and public health experts in survey methodology and vaccine attitudes. The survey was pilot tested with our institution’s interdisciplinary health services Program on Women’s Health Effectiveness Research. The questionnaire is available online.^11^

Our primary outcome was receipt of or intent to receive the COVID-19 vaccine (“What have you done or are you planning to do regarding the COVID-19 vaccine”), measured using the following four answer categories: “I have received or am scheduled to receive the vaccine already,” “ I intend to get it as soon as I can,” “I do not intend to get it soon but might sometime in the future,” or “I do not intend to ever get the vaccine.” Other key outcomes included a series of items assessing potential reasons for hesitancy or concerns about the COVID-19 vaccine.

Personal demographic characteristics of participants were captured using standard items to assess age, race, and ethnicity. Gender response options included “male,” “female,” “trans male/trans man,” “trans female/trans woman,” “gender queer/gender non-conforming,” and “different identity.” Personal history of COVID-19 infection was captured by creation of a binary experience variable if the respondent answered yes to either of the following statements: “I believe that I have already had COVID-19” and “I have been hospitalized with COVID-19.”

To explore occupational variations within the healthcare system, we asked each respondent to self-categorize their occupational role according to the following eight categories: 1) nurse; 2) nurse practitioner/nurse midwife/physician assistant; 3) physician; 4) other staff providing clinical care (e.g., phlebotomist, radiology technician); 5) other staff not providing clinical care but with clinical contact (e.g., unit clerk, food service staff); 6) trainee (i.e., medical/graduate/post-doctoral student); 7) administrative or research staff and faculty with no clinical contact; 8) all other employees. We also separately asked whether respondents’ job responsibilities required any time on high-risk COVID-19 units (i.e., the Emergency Department, Intensive Care Unit, the Birth Center, or Trauma Bay or Operating Rooms).

### Statistical Analysis

The primary outcome of this study was receipt of or intention to receive the COVID-19 vaccine. We conducted descriptive analyses to ascertain the proportion of individuals with each vaccine receipt/intention category, overall and by sociodemographic and occupational characteristic groups. We combined individuals responding “I have received or am scheduled to receive the vaccine already” or “I intend to get it as soon as I can” into one group (“Received/Scheduled/ASAP”) and described demographic characteristics across three vaccine intention groups: 1) Received/Scheduled/ASAP, 2) Not now, and 3) Not ever. We then described vaccine intention by employee role stratified by both gender and race/ethnicity. We collapsed gender into “female,” “male,” and “other;” some exhibits include only female and male groups, due to small numbers. Due to small numbers of respondents in certain racial and ethnic groups, we collapsed race/ethnicity into four categories (Hispanic, non-Hispanic White, non-Hispanic Asian, and non-Hispanic Black/Mixed/Other). This last category was created because when independently assessed, the groups all behaved similarly. Chi Square test was used to carry out significant difference tests and p values <0.05 were considered statistically significant.

We constructed a multivariable ordinal logistic regression model to determine demographic factors independently associated COVID-19 vaccine receipt/intention. We used SAS 9.4 (Cary, NC) to perform the statistical analyses. The study was deemed exempt research by the study site’s Institutional Review Board (HUM00193484). Participation was voluntary and consent was attained through survey continuation.

## RESULTS

A total of 39,259 individual and group email accounts received the study invitation and 11,387 individuals responded. This corresponds to a response rate of at least 29% of the employee population (the presence of group email accounts in the distribution list means that this calculation is an underestimate of the true response rate). Mirroring the demographics of the health system employee population,^12^ participants were largely non-Hispanic White, evenly distributed across age groups, and reflective of a wide variety of professional roles within the healthcare system (Table 1).

**Table 1.** Sociodemographic and occupational characteristics of respondents by vaccine intention.

| Characteristic | Vaccine Intention |  |  | p value<sup>§</sup> |
| --- | --- | --- | --- | --- |
|  | Received/Scheduled/<br>ASAP*<br>(n=9627) | Not Now<sup>†</sup><br>(n=954) | Not Ever<sup>‡</sup><br>(n=369) |  |
| Role |  |  |  | <0.001 |
| Nurse | 1463 (87.4) | 143 (8.5) | 68 (4.1) |  |
| NP/Nurse Midwife/PA | 339 (93.7) | 18 (5.0) | 5 (1.4) |  |
| Physician | 1138 (99.5) | 4 (0.4) | 2 (0.2) |  |
| Other staff providing clinical care | 1460 (82.3) | 222 (12.5) | 92 (5.2) |  |
| Other staff not providing clinical care but with clinical contact<sup>¶</sup> | 1138 (87.9) | 117 (9.0) | 39 (3.0) |  |
| Trainee<sup>#</sup> | 350 (97.2) | 8 (2.2) | 2 (0.6) |  |
| Admin/Research Staff/Faculty with no clinical contact | 2447 (88.2) | 248 (8.9) | 80 (2.9) |  |
| All other employees | 1006 (85.3) | 116 (9.8) | 58 (4.9) |  |
| High Risk Unit** |  |  |  | 0.04 |
| Yes | 2757(89.1) | 243 (7.9) | 92 (3.0) |  |
| No | 6870 (87.4) | 711 (9.1) | 277 (3.5) |  |
| Personal Infection with COVID<sup>††</sup> |  |  |  | <0.001 |
| Yes | 1059 (73.8) | 259 (18.0) | 118 (8.2) |  |
| No | 8568 (90.1) | 695 (7.3) | 251 (2.6) |  |
| Gender |  |  |  | <0.001 |
| Female | 7137 (86.9) | 791 (9.6) | 287 (3.5) |  |
| Male | 2100 (94.1) | 85 (3.8) | 47 (2.1) |  |
| Other | 72 (86.8) | 6 (7.2) | 5 (6.0) |  |
| Race/Ethnicity |  |  |  | <0.001 |
| Hispanic | 301 (88.0) | 28 (8.2) | 13 (3.8) |  |
| Non-Hispanic Asian | 653 (96.2) | 23 (3.4) | 3 (0.4) |  |
| Non-Hispanic Black/Mixed/Other | 622 (77.0) | 132 (16.3) | 54 (6.7) |  |
| Non-Hispanic White | 7668 (89.1) | 674 (7.8) | 261 (3.0) |  |
| Age, years |  |  |  | <0.001 |
| ≤34 | 2483 (84.6) | 333 (11.4) | 118 (4.0) |  |
| 35-44 | 2248 (87.3) | 231 (9.0) | 97 (3.8) |  |
| 45-54 | 2065 (88.0) | 200 (8.5) | 81 (3.5) |  |
| 55-64 | 2009 (92.5) | 115 (5.3) | 47 (2.2) |  |
| ≥65 | 562 (97.2) | 12 (2.1) | 4 (0.7) |  |

\*I have received or am scheduled to receive the vaccine already, or I intend to get it as soon as I can
<sup>†</sup>I do not intend to get the vaccine soon but might get it in the future
<sup>‡</sup>I do not intend to ever get the vaccine
<sup>§</sup>P values generated via Chi Square and Fisher Exact tests
<sup>||</sup>Phlebotomist, radiology/imaging technician, medical assistant, etc.
<sup>¶</sup>Unit clerk, check-in/check-out, food service, etc.
<sup>#</sup>Includes medical student, graduate student, post-doctoral student
<sup>\*\*</sup>Emergency Department, Intensive Care Unit, the Birth Center, or Trauma Bay or Operating Rooms
<sup>††</sup>Includes "I believe that I have already had COVID-19" and "I have been hospitalized with COVID-19"
Data presented as n (%). Proportions may not round to 100 due to rounding.
ASAP, as soon as possible; NP, Nurse Practitioner; PA, Physician Assistant

As of the date of survey response, most respondents had received or were scheduled to receive the COVID-19 vaccine (n=9081, 79.8%) or planned to receive the COVID-19 vaccine as soon as possible (n=546, 4.8%). However, more than 11% of respondents were vaccine hesitant, either not intending to get it soon but might in the future (n=954, 8.4%) or not intending to ever get the vaccine (n=369, 3.2%). Vaccine intention differed significantly across professional role, high-risk unit, personal infection with COVID-19, gender, race/ethnicity, and age (Table 1). In univariate analysis, personal infection with COVID-19, female gender, non-Hispanic Black/Mixed/Other race/ethnicity, and younger age were significantly associated with decreased vaccination intentions.

Vaccine intentions also demonstrated notable shifts across professional roles, especially when stratified by gender (Figure 1) and race/ethnicity (Figure 2). Physicians, nurse practitioners/midwives/physician assistants, and trainees demonstrated the highest vaccine acceptance, while nurses, other staff providing clinical care, those without clinical responsibilities but with clinical contact, and those with other professional roles not involving clinical contact demonstrated lower vaccine acceptance. In these professional groups with lower vaccine acceptance, there was a pattern of greater vaccine hesitancy among females, who demonstrated higher proportions of “not now” and “not ever” intentions compared to males (Figure 1). When employment roles were stratified by race/ethnicity (Figure 2), physicians had extremely high vaccine acceptance across all racial groups, with only five physicians in the whole sample reporting vaccine hesitancy. There were no Non-Hispanic Black/Mixed/Other physicians who reported vaccine hesitancy. In contrast, in nearly all other employment groups, the highest rates of vaccine hesitancy were observed among Non-Hispanic Black/Mixed/Other employees. For example, “not now” and “not ever” intentions were reported by more than 1 in 4 other staff providing clinical care (“not now,” 22.8%; “not ever,” 12.6%) and staff not providing clinical care but with clinical contact (“not now,” 20.0%; “not ever,” 6.4%). Employees who self-identified as Hispanic also demonstrated patterns of higher vaccine hesitancy, with 19% of Hispanic nurses reporting “not now” and 8.7% reporting “not ever” vaccine intentions. Across all employee groups (with the exception of physicians, who demonstrated globally high rates of acceptance), employees who self-identified as Asian had the highest rates of vaccine acceptance compared to all other racial groups. Formal statistical testing for interaction between gender groups and role and racial-ethnic groups and role could not be reliably performed due to small overall numbers in some subgroups.

**Figure 1.**
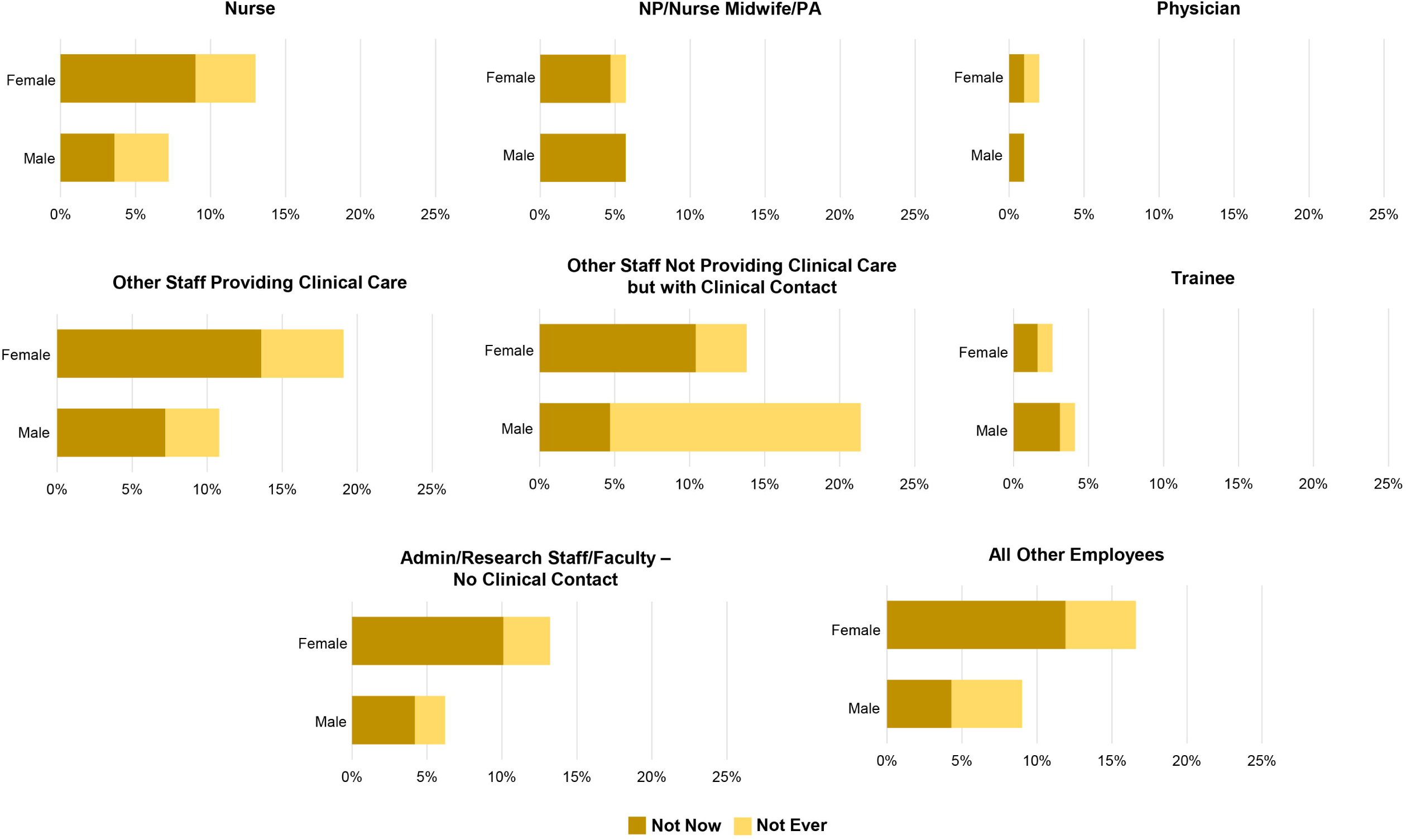
Not Now and Not Ever vaccine intentions according to medical center employee role stratified by gender. *Not Now*=I do not intend to get the vaccine soon but might get it in the future; *Not Ever*=I do not intend to ever get the vaccine; *Other Staff Providing Clinical Care*=phlebotomist, radiology/imaging technician, medical assistant, etc.; *Other Staff Not Providing Clinical Care but with Clinical Contact*=unit clerk, check-in/check-out, food service, etc.; *Trainee*=medical student, graduate student, post-doctoral student; *NP*=Nurse Practitioner; *PA*=Physician Assistant

**Figure 2.**
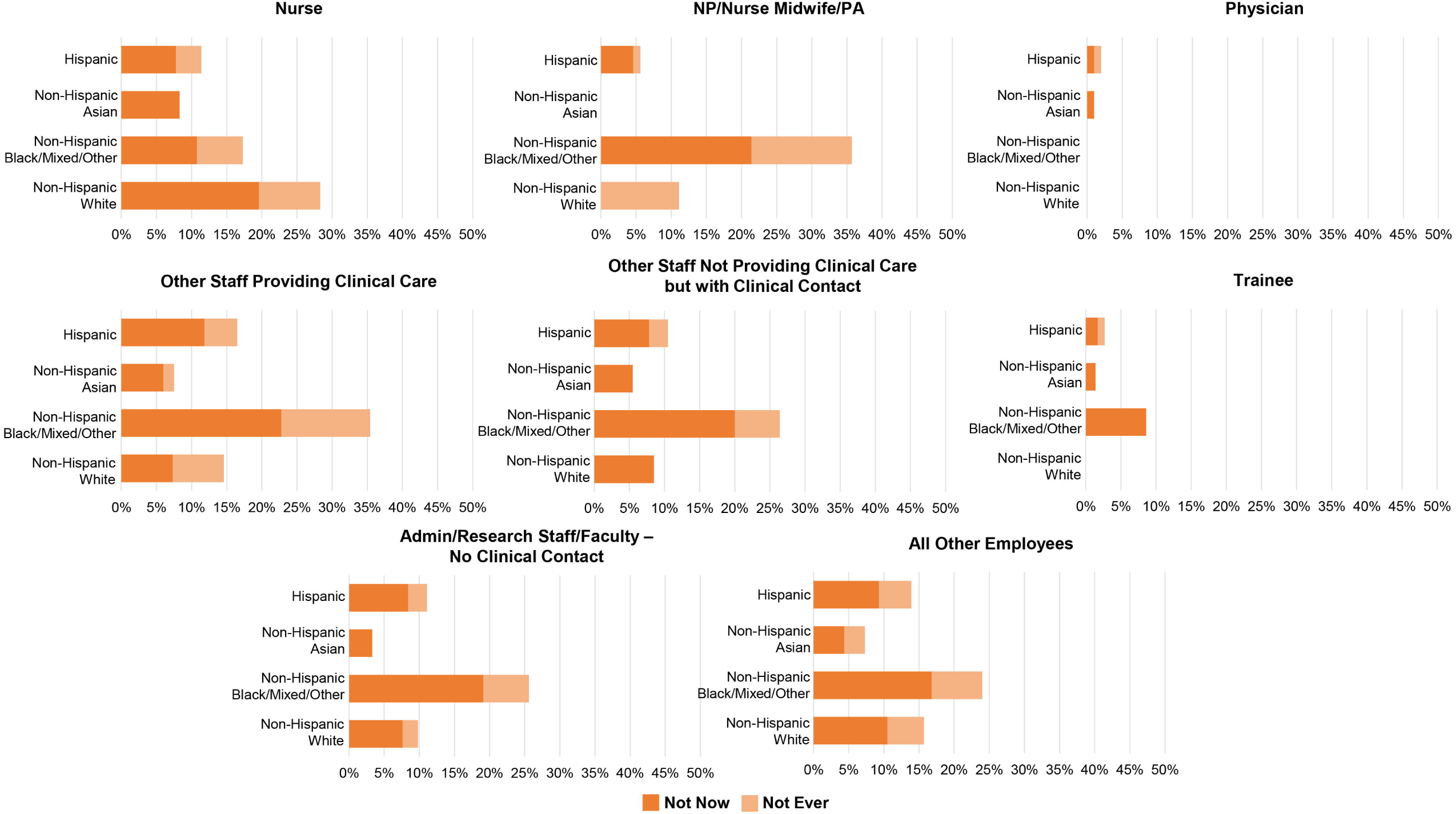
Not Now and Not Ever vaccine intentions according to medical center employee role stratified by race/ethnicity. *Not Now*=I do not intend to get the vaccine soon but might get it in the future; *Not Ever*=I do not intend to ever get the vaccine; *Other Staff Providing Clinical Care*=phlebotomist, radiology/imaging technician, medical assistant, etc.; *Other Staff Not Providing Clinical Care but with Clinical Contact*=unit clerk, check-in/check-out, food service, etc.; *Trainee*=medical student, graduate student, post-doctoral student; *NP*=Nurse Practitioner; *PA*=Physician Assistant

In multivariable logistic regression models predicting vaccine receipt/intention, there were strong associations with professional role (Table 2). Compared to nurses, physicians (aOR 22.2, 9.1-54.3), trainees (aOR 5.9, 3.0-11.4), and nurse practitioners/nurse midwives/physician assistants (aOR 1.9, 1.2-3.0) were significantly more likely to demonstrate vaccine acceptance, whereas other clinical staff were significantly less likely (aOR 0.8, 0.6-0.9) and all other employees marginally less likely (aOR 0.8, 0.6-1.0). Working in a high-risk clinical unit versus not was marginally associated with a higher rate of vaccine acceptance (aOR 1.2, 1.0-1.4) after controlling for other covariates. Prior infection with COVID-19 was associated with significantly lower vaccine acceptance (aOR 0.4, 0.3-0.4). Male employees were almost twice as likely to accept vaccines compared to female employees (aOR 1.9, 1.6-2.4). Compared to non-Hispanic White respondents, non-Hispanic Asian respondents had significantly higher odds of vaccine acceptance (aOR 2.3, 1.5-3.4), while non-Hispanic Black/Mixed/Other respondents had significantly lower odds of vaccine acceptance after controlling for other covariates (aOR 0.4, 0.4-0.5). Increasing age was significantly associated with vaccine acceptance, with increased acceptance across age groups compared to the reference group of respondents ≤34 years of age. In subgroup analyses, associations between sociodemographic and occupational characteristics and vaccine acceptance persisted among only female respondents, but diverged among only male respondents. Specifically, compared to male nurses, only male physicians had higher odds of vaccine acceptance, while other roles all demonstrated statistically similar odds. Respondents with prior infection with COVID-19 and non-Hispanic Black/Mixed/Other respondents continued to demonstrate decreased odds compared to peer groups, but age was no longer strongly predictive of vaccine intention.

**Table 2.** Logistic regression model predicting vaccine intention, overall, and within female and male subgroups.

| Covariate | Whole Cohort<br>Odds Ratio (95%CI) | Female Subgroup<br>Odds Ratio (95%CI) | Male Subgroup<br>Odds Ratio (95%CI) |
| --- | --- | --- | --- |
| Role |  |  |  |
| Nurse | Ref | Ref | Ref |
| Physician | 22.2 (9.1-54.3) | 26.7 (8.5-83.9) | 13.9 (2.9-65.9) |
| NP/nurse midwife/PA | 1.9 (1.2-3.0) | 2.3 (1.4-3.7) | 1.0 (0.2-5.2) |
| Other staff providing clinical care* | 0.8 (0.6-0.9) | 0.7 (0.6-0.9) | 0.6 (0.3-1.4) |
| Other staff without clinical responsibilities but with clinical contact<sup>†</sup> | 1.1 (0.9-1.4) | 1.0 (0.8-1.3) | 1.1 (0.5-2.6) |
| Trainee<sup>‡</sup> | 5.9 (3.0-11.4) | 9.8 (3.9-24.3) | 1.9 (0.5-6.9) |
| Admin/Research Staff/Faculty with no clinical contact | 1.0 (0.8-1.3) | 1.0 (0.8-1.3) | 0.9 (0.4-2.0) |
| All other employees | 0.8 (0.6-1.0) | 0.8 (0.6-1.0) | 0.6 (0.3-1.4) |
| High Risk Unit |  |  |  |
| No | Ref | Ref | Ref |
| Yes | 1.2 (1.0-1.4) | 1.3 (1.0-1.5) | 1.0 (0.6-1.6) |
| Prior infection with COVID-19 |  |  |  |
| No | Ref | Ref | Ref |
| Yes | 0.4 (0.3-0.4) | 0.4 (0.3-0.4) | 0.3 (0.2-0.5) |
| Gender |  |  |  |
| Female | Ref | N/A | N/A |
| Male | 1.9 (1.6-2.4) | N/A | N/A |
| Other | 1.5 (0.8-3.1) | N/A | N/A |
| Race/Ethnicity |  |  |  |
| Non-Hispanic White | Ref | Ref | Ref |
| Non-Hispanic Asian | 2.3 (1.5-3.4) | 2.6 (1.6-4.1) | 1.2 (0.5-2.9) |
| Non-Hispanic Black/Mixed/Other | 0.4 (0.4-0.5) | 0.5 (0.4-0.6) | 0.3 (0.2-0.4) |
| Hispanic | 1.0 (0.7-1.4) | 1.0 (0.7-1.5) | 1.1 (0.3-3.8) |
| Age, years |  |  |  |
| ≤34 | Ref | Ref | Ref |
| 35-44 | 1.3 (1.1-1.5) | 1.3 (1.1-1.6) | 1.2 (0.7-2.0) |
| 45-54 | 1.6 (1.3-1.9) | 1.6 (1.4-2.0) | 1.3 (0.7-2.2) |
| 55-64 | 2.5 (2.1-3.1) | 2.7 (2.1-3.3) | 2.0 (1.1-3.7) |
| 65+ | 5.3 (3.1-9.0) | 6.2 (3.3-11.4) | 4.0 (1.2-13.9) |

\*Phlebotomist, radiology/imaging technician, medical assistant, etc.
<sup>†</sup>Unit clerk, check-in/check-out, food service, etc.
<sup>‡</sup>Includes medical student, graduate student, post-doctoral student NP, Nurse Practitioner; PA, Physician Assistant

Overall, 29.6% of respondents reported at least one concern about COVID-19 vaccination, with vaccine concerns varying by vaccine intention (Table 3). Among those with vaccine hesitancy, the most commonly reported concerns were mistrust in the vaccine because of how quickly it was developed, insufficient safety and effectiveness data, disbelief that the vaccine would protect against COVID-19 infection, and concerns about serious side effects from vaccination. Even in respondents accepting the COVID-19 vaccine, a notable proportion also harbored vaccine concerns, with 21.2% responding affirmatively to at least one concern, including 6% concerned about how quickly the vaccine was developed, 6% concerned about lack of safety and effectiveness data, and 7% concerned about the potential for bad reactions to the vaccine.

**Table 3.**
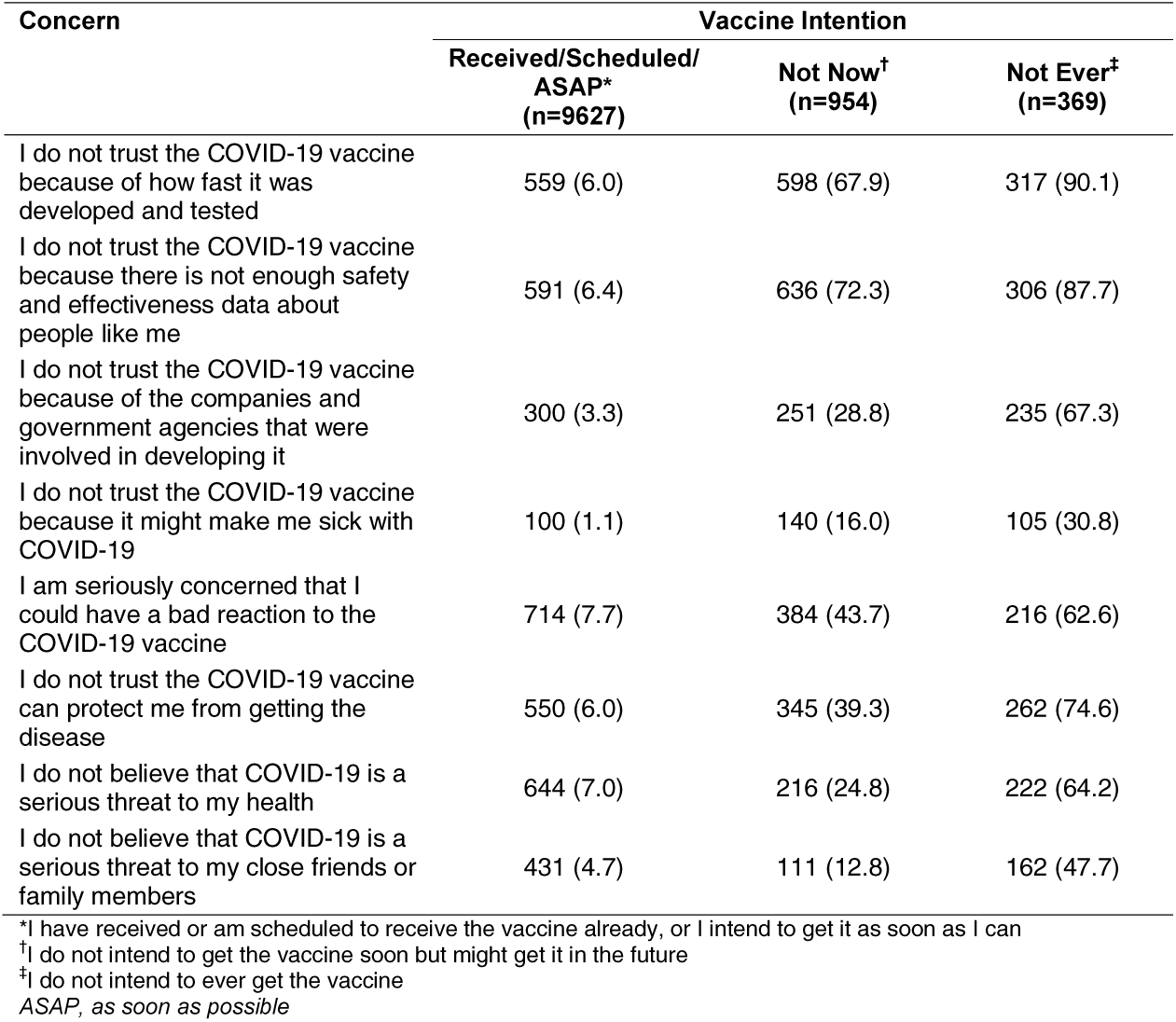
Concerns about the COVID-19 vaccine by vaccine intention group.

| Concern | Vaccine Intention |  |  |
| --- | --- | --- | --- |
|  | Received/Scheduled/<br>ASAP*<br>(n=9627) | Not Now<sup>†</sup><br>(n=954) | Not Ever<sup>‡</sup><br>(n=369) |
| I do not trust the COVID-19 vaccine because of how fast it was developed and tested | 559 (6.0) | 598 (67.9) | 317 (90.1) |
| I do not trust the COVID-19 vaccine because there is not enough safety and effectiveness data about people like me | 591 (6.4) | 636 (72.3) | 306 (87.7) |
| I do not trust the COVID-19 vaccine because of the companies and government agencies that were involved in developing it | 300 (3.3) | 251 (28.8) | 235 (67.3) |
| I do not trust the COVID-19 vaccine because it might make me sick with COVID-19 | 100 (1.1) | 140 (16.0) | 105 (30.8) |
| I am seriously concerned that I could have a bad reaction to the COVID-19 vaccine | 714 (7.7) | 384 (43.7) | 216 (62.6) |
| I do not trust the COVID-19 vaccine can protect me from getting the disease | 550 (6.0) | 345 (39.3) | 262 (74.6) |
| I do not believe that COVID-19 is a serious threat to my health | 644 (7.0) | 216 (24.8) | 222 (64.2) |
| I do not believe that COVID-19 is a serious threat to my close friends or family members | 431 (4.7) | 111 (12.8) | 162 (47.7) |

\*I have received or am scheduled to receive the vaccine already, or I intend to get it as soon as I can
<sup>†</sup>I do not intend to get the vaccine soon but might get it in the future
<sup>‡</sup>I do not intend to ever get the vaccine ASAP, as soon as possible

## DISCUSSION

During a pandemic, health systems must rapidly respond to keep critical infrastructure operating. Employee vaccination is central to ensuring a healthy workforce, but vaccine concerns may limit widespread uptake of vaccines. In a timely assessment of over 11,000 medical center employees’ COVID-19 vaccine intentions conducted just as vaccination was occurring within a healthcare system, we document important patterns in COVID-19 vaccine acceptance and hesitancy. Despite high vaccine acceptance overall, there was notable variation across professional roles, with physicians, nurse practitioners/nurse midwives/physician assistants, and medical trainees demonstrating significantly higher vaccine acceptance than nurses, other staff with clinical roles, other staff without clinical responsibilities but with clinical contact, and non-clinical faculty and staff. Women, individuals identifying as non-Hispanic Black/Mixed/Other, those with prior COVID-19 infection, and younger adults were also significantly less likely to have accepted COVID-19 vaccination.

Prior studies, conducted before Phase III clinical trial results were released and FDA EUA was granted, hinted at discrepancies in hypothetical COVID-19 vaccine acceptability by healthcare worker role, with higher hesitancy among non-physician personnel.^13,14^ Another recent study documented higher vaccine acceptance among clinical compared to non-clinical health system workers.^15^ Our study significantly advances this prior work by newly documenting real-world COVID-19 vaccine acceptance among medical center employees and significant variation in vaccine delay/deferral across professional role and demographic characteristics—especially gender, race/ethnicity, and age. Our findings call for nuanced, nimble communication strategies that account for the multiple intersecting characteristics that may affect a medical center employee’s risk of vaccine acceptance, recognizing, as others have noted, that no demographic group is a monolithic group with a single, collective viewpoint.^16,17^

Our findings advance other studies suggesting that vaccine hesitancy among many healthcare workers is less driven by overall vaccine skepticism and more by specific concerns about the lack of long-term data on safety, efficacy, and potential side effects of the COVID-19 vaccines.^13,14,18,19^ Many concerns were cross-cutting, noted even among some of those accepting COVID-19 vaccination. These concerns constitute potential targets for communication interventions to reduce vaccine concerns. Clear, consistent messaging about the rigorous clinical trials process, ongoing safety data, and FDA approval process may help mitigate hesitancy. However, decades of anthropological research caution against simply providing “more and better” information without also leveraging empathy.^20,21^ Acknowledging the legitimacy of vaccine concerns and fear of the unknown may be a critical tool to help dismantle vaccine hesitancy. In fact, as Lisa Rosenbaum has suggested,^21^ healthcare workers with vaccine concerns may be potent vaccine champions, uniquely poised to understand patients who are reluctant to get vaccinated, to communicate that what is known about the vaccine is more important than what is unknown, and to change hearts and minds to increase vaccine uptake.

Our findings hold important implications for clinical practice, public health efforts, health policy, and research, both immediately during the COVID-19 pandemic and more broadly to guide efforts in future infectious disease outbreaks. More robust communication efforts are needed to address vaccine concerns among medical center employees, particularly among non-physician groups, women, and non-Hispanic Black/Mixed/Other individuals. Increasing vaccine confidence among healthcare workers is foundational to successfully instilling vaccine acceptance in other populations and achieving vaccine coverage sufficient to stop viral spread.^22,23^ Emerging research suggests that “Be a protector” messaging, which emphasizes empathy and being a protector for loved ones, may better reduce vaccine resistance than traditional “get the vaccine” messages, which may fuel resistance by eliciting shame and anger about being told what to do.^24^ “Protector” schema messages may be particularly salient for healthcare workers, who have a shared goal and obligation to protect their patients and community. Tailored messaging that targets the informational needs and preferences of specific groups may also be helpful.

### THE CONVERSATION

*Between Us, About Us*, for example, is a novel, living video library featuring Black doctors, nurses, and researchers aiming to provide Black communities with credible information about COVID-19 vaccination.^25^

A key strength of this study is a large, real-world sample of medical center employees drawn from the entire population of an academic medical center. Moreover, we measured respondents’ self-reported vaccine acceptance or deferral/declination at the time the vaccine was available to them and when the initial scientific efficacy data was available, whereas many prior studies have evaluated anticipated intentions regarding hypothetical vaccines. Study limitations include an opt-in sampling approach, which may bias toward those with stronger opinions (either positive or negative) about the COVID-19 vaccine, and our cross-sectional design, which cannot capture rapidly evolving attitudes or beliefs.

In this large study of a diverse sample of medical center employees’ COVID-19 vaccine intentions and attitudes as vaccination was made available to them, we document high vaccine acceptance overall but notable variation across health care employee roles, gender, and racial-ethnic groups. Our findings suggest important opportunities to empathetically engage those with COVID-19 vaccine concerns and optimize vaccine coverage across our healthcare system.

## Supporting information

CHERRIES reporting checklist for online surveys

Author COI disclosure forms

## Data Availability

Data available upon request from the first author.

## Contributors

The authors thank Sarah Block for tireless assistance with manuscript preparation.

## Funding

None

## Prior presentations

None

## Notes

### Competing Interest Statement

The authors have declared no competing interest.

### Funding Statement

No funding was received for the presented work.

### Author Declarations

University of Michigan Institutional Review Board (HUM00193484)

